# Efficacy of Repetitive Transcranial Magnetic Stimulation in Alzheimer’s Disease: A Systematic Review and Meta-Analysis of Cognitive, Mood, and Functional Outcomes

**DOI:** 10.1101/2025.08.31.25334781

**Authors:** Bavurothu Sharanya Kumar, Anwar Syed, Ekta, Aishwarya Raparthi, Abhishek Palaniappan, Shradha Pandurang Kakde, M N Shreelakshmi, Shankar Biswas, Sheikh Ramiz Ahmed, Rakhshanda Khan, Harshawardhan Dhanraj Ramteke

**Affiliations:** Andhra medical college, AP, India; Bhaskar medical college, Moinabad, India; Christian medical college, India; KMCH medical college, India; MGM Medical college and research center, India; Mandya institute of medical sciences, Mandya, Karnataka, India; Ivano-Frankivsk national medical university, Ukraine; Government medical college kolkata, Kolkata India; Ayaan institute of medical sciences, Moinabad, Ranga Reddy, India; Anhui medical university, Hefei, China

**Author notes:** **Corresponding author** Rakhshanda khan Ayaan institute of medical sciences, Moinabad, Ranga reddy, India.

**Keywords:** Alzheimer’s disease, repetitive transcranial magnetic stimulation, cognitive function, systematic review, meta-analysis, MMSE, ADAS-cog, mood, functional performance, adverse events

## Abstract

**Introduction:** Alzheimer’s disease (AD) is a neurodegenerative disorder characterized by cognitive decline and changes in mood and behavior. While current pharmacological treatments offer limited symptom relief, non-invasive therapies like repetitive transcranial magnetic stimulation (rTMS) have emerged as potential alternatives. This systematic review and meta-analysis aimed to assess the effects of rTMS on cognitive function, mood, and functional abilities in AD patients.

**Methods:** A comprehensive search of PubMed, Embase, and Cochrane databases was conducted, including studies published until December 2024. Randomized controlled trials (RCTs) involving AD patients treated with rTMS were included, with cognitive outcomes measured using the Mini-Mental State Examination (MMSE) and Alzheimer’s Disease Assessment Scale-Cognitive Subscale (ADAS-cog). Secondary outcomes included mood (Geriatric Depression Scale, GDS), global impression (Clinician’s Global Impression of Change, CGIC), and functional performance (Instrumental Activities of Daily Living, IADL). Data analysis was conducted using Stata 18.0.

**Results:** Thirty-five studies with 1794 participants (887 males, 869 females) were included. The overall mean difference in cognition (MMSE) was -0.48 (95% CI [-1.41, 0.46], I² = 99.42%), suggesting no significant cognitive improvement. In the ADAS-cog scale, the overall mean difference was -2.94 (95% CI [-5.13, -0.75], I² = 98.61%), indicating a significant treatment effect in favor of rTMS. For mood and functional performance, no significant difference was observed (mean difference -0.96, 95% CI [-6.21, 4.30], I² = 97.38%). The incidence of adverse events, including headaches and scalp pain, showed no significant differences between groups (log risk ratio for headaches: -0.31, 95% CI [-0.84, 0.22], I² = 3.20%).

**Conclusion:** While rTMS shows potential for improving cognition in AD patients, its effects remain inconsistent. Significant heterogeneity across studies underscores the need for standardized protocols in future research. Additionally, rTMS appears safe, with no significant increase in adverse events.

## Introduction

Alzheimer’s disease (AD) is a progressive neurodegenerative disorder that leads to severe impairments in cognitive functions, such as memory, language, and executive function, as well as notable changes in mood, personality, and behavior. As the disease progresses, individuals face increasing difficulties with daily activities, creating a significant burden on caregivers, healthcare systems, and society as a whole [1]. The current pharmacological treatments, such as cholinesterase inhibitors (Donepezil, Rivastigmine) and glutamate regulators (Memantine), provide modest improvements in cognitive symptoms but are not disease-modifying, underscoring the need for alternative therapeutic strategies [2].

One promising approach is repetitive transcranial magnetic stimulation (rTMS), a non-invasive brain stimulation technique that modulates neural activity in targeted brain regions by using electromagnetic induction. By applying a magnetic coil to the scalp, rTMS generates electrical currents that can either excite or inhibit cortical neurons, depending on the frequency of stimulation [3]. High-frequency rTMS has been shown to enhance neural activity, while low-frequency stimulation can suppress it [4]. Furthermore, rTMS has the potential to influence broader brain networks beyond the primary stimulation site, possibly inducing long-lasting synaptic changes that may contribute to its therapeutic effects [5]. Given these mechanisms, rTMS has been investigated for treating various neurological and psychiatric disorders, including stroke, Parkinson’s disease, depression, and schizophrenia [6].

Recent clinical trials have explored the safety and efficacy of rTMS in AD patients, with several studies suggesting that it may have beneficial effects on cognitive function [7,8]. However, the findings remain inconsistent due to variations in study design, sample size, and outcome measures, making it difficult to reach a definitive conclusion [9]. Notably, prior systematic reviews have primarily focused on cognitive outcomes, without sufficiently addressing the impact of rTMS on mood, overall clinical impression, or functional performance, all of which are critical aspects of AD management [10,11]. To fill this gap, we present a comprehensive systematic review and meta-analysis of recent randomized controlled trials (RCTs) to evaluate the effects of rTMS on cognition, mood, global impression, and functional abilities in individuals with Alzheimer’s disease.

## Methods

### Search Strategy

A comprehensive search was carried out in the PubMed, Embase, and Cochrane databases to identify relevant studies published up to December 31, 2024, without language restrictions. Key search terms included "transcranial magnetic stimulation," "Magnetic Stimulation, Transcranial," "Alzheimer’s disease," "Alzheimer’s Dementia," and "Alzheimer Dementia." In addition, reference lists from pertinent articles were manually reviewed to identify any further studies that might be relevant. This process followed the PRISMA (Preferred Reporting Items for Systematic Reviews and Meta-Analyses) guidelines [12], ensuring thorough and transparent search methods. The Protocol was registered with PROSPERO (CRD420251137461).

### Study Selection and Data Extraction

Eligibility for inclusion was based on the following criteria: (1) Participants had a clinical diagnosis of Alzheimer’s disease; (2) Repetitive transcranial magnetic stimulation (rTMS) was administered either alone or in combination with other treatments; (3) A sham rTMS group served as a control; (4) Cognitive function was assessed using standard measures such as the Alzheimer’s Disease Assessment Scale-Cognitive Subscale (ADAS-cog) or the Mini-Mental State Examination (MMSE); (5) The study was a randomized controlled trial (RCT).

Data extraction was conducted independently by two authors. Information collected included participant demographics (total number, age, gender, education level, baseline MMSE scores), rTMS parameters (target brain regions, stimulation frequency, intensity, number of sessions, pulses per session, and details regarding sham stimulation), outcome measures, follow-up duration, and any reported adverse effects. The primary outcome of interest was cognitive function, evaluated using either ADAS-cog or MMSE. Secondary outcomes included mood (measured by the Geriatric Depression Scale, GDS), overall clinical impression (Clinician’s Global Impression of Change, CGIC), and functional performance (assessed using the Instrumental Activities of Daily Living, IADL scale).

For the meta-analysis, both baseline and post-treatment outcome data were gathered. If studies provided data at multiple time points, the immediate post-treatment values were prioritized. When necessary, data were not available, corresponding authors were contacted to request the missing information.

### Risk of Bias Assessment

The risk of bias in the included studies was evaluated using the Cochrane Collaboration’s Risk of Bias Tool (ROB 2.0) [13]. Disagreements regarding risk of bias were resolved through discussion or consultation with an independent third party. The tool evaluates bias across several domains, including random sequence generation, allocation concealment, participant and personnel blinding, blinding of outcome assessment, completeness of outcome data, selective reporting of results, and any other sources of bias. Each domain was rated as having a low, high, or unclear risk of bias.

### Statistical Analysis

Statistical analyses were conducted using Stata 18.0. For ordinal rating scales, data were treated as continuous variables, with mean changes from baseline reported. The treatment effect was represented as the mean difference (MD) with 95% confidence intervals (CI). When standard deviations (SD) were not provided, these were calculated from p-values, in line with the guidelines from the Cochrane Handbook for Systematic Reviews of Interventions.

To assess heterogeneity, both the Chi² test and the I² statistic were employed. A significance threshold of P < 0.10 was used for the Chi² test, while an I² value greater than 50% indicated substantial heterogeneity. In cases of low heterogeneity (I² < 50%), a fixed-effects model was used to calculate pooled estimates of mean differences between the intervention groups. If significant heterogeneity was present, a random-effects model was applied.

Subgroup analysis was performed based on rTMS stimulation frequency and targeted brain regions. Sensitivity analysis using a leave-one-out approach was conducted to assess the influence of individual studies on the overall results in the presence of significant heterogeneity. Due to the inclusion of fewer than 10 studies, publication bias was not assessed.

## Results

### Demographics

A total of 1,274 studies were identified across five databases. After removing 637 duplicates and excluding 256 ineligible records, 77 additional records were removed for other reasons. This left 304 records for screening of which 213 were excluded. The reports sought for retrieval were 91of which 41 could not be resolved. The final 50 were assessed for eligibility, of which 35 were analyzed [14–48]. Figure 1 has the PRISMA flow diagram. The total sample of the population was 1794, male patients were 887, whereas female patients were 869, the average age was 69.8 ± 12 years and average follow up time in months was 5.4 ± 2.5. All of the details are given in Table S1.

**Figure 1.**
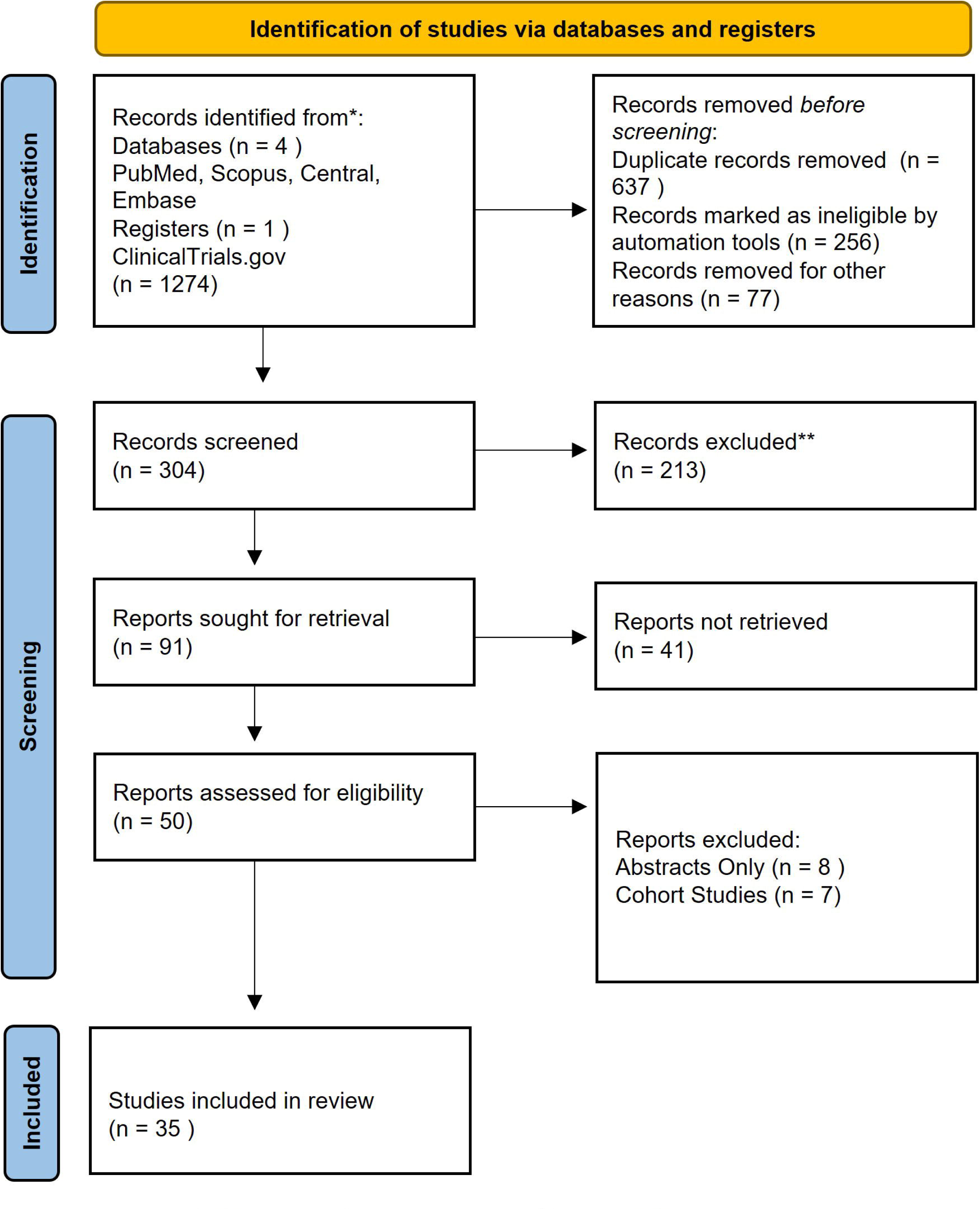
Prisma Flow Diagram

### Cognition after repetitive transcranial magnetic stimulation (rTMS) versus sham rTMS in MMSE Scale

The overall mean difference between treatment and control groups is -0.48 with a 95% CI of [-1.41, 0.46], suggesting a small negative effect of the treatment compared to the control, though the confidence interval includes 0, indicating a lack of significant difference between groups. The heterogeneity statistics indicate a high variability in effect sizes across studies (I² = 99.42%), implying that the results from different studies may not be directly comparable and could be influenced by differing methodologies, populations, or interventions.

Notably, studies like "Ahmed et al. 2012" and "Koch et al. 2025" demonstrate more substantial effects with wider confidence intervals, while others, such as "Zhao et al. 2017" and "Leocani et al. 2012," show smaller effects and tighter intervals. This variance may be due to differences in sample characteristics, such as disease severity or treatment regimens. The overall analysis is based on a random-effects model, suggesting that the true effect of the treatment could vary across different studies, which may influence the interpretation of the intervention’s efficacy. Figure 2.

**Figure 2.**
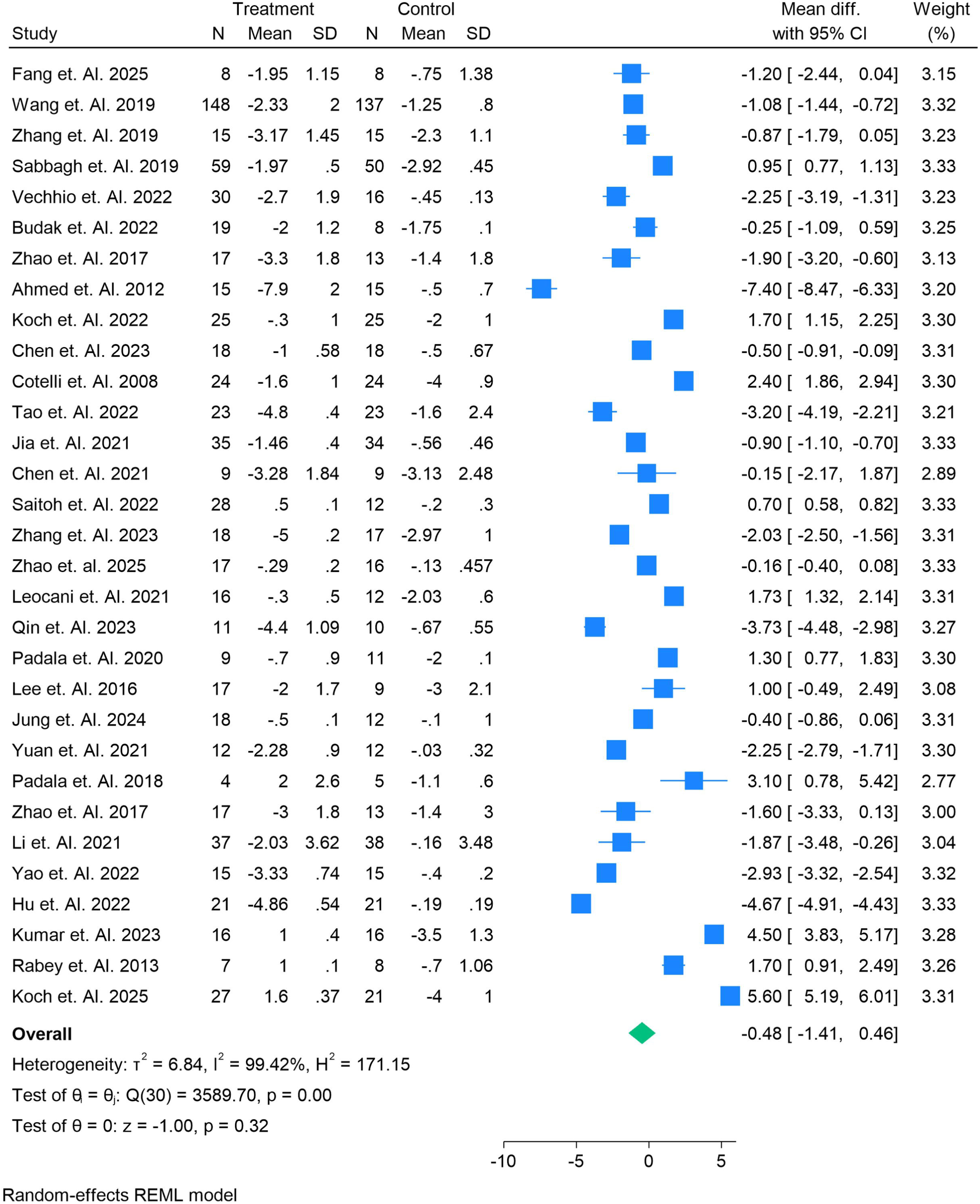
Meta-analysis of cognition after repetitive transcranial magnetic stimulation (rTMS) versus sham rTMS in MMSE Scale.

### Cognition after repetitive transcranial magnetic stimulation (rTMS) versus sham rTMS in ADAS-cog Scale

The analysis reports the mean difference between the two groups, with 95% confidence intervals (CIs) shown for each study. The overall mean difference is -2.94, with a 95% CI of [-5.13, -0.75], indicating a significant treatment effect favoring the treatment group. The heterogeneity statistics (I² = 98.61%) indicate considerable variability between the studies, suggesting that differences in study design, populations, or treatment protocols may contribute to this variation.

In this analysis, studies such as "Wang et al. 2019" and "Li et al. 2021" contribute more weight due to their larger sample sizes, and they show relatively smaller mean differences compared to other studies, such as "Chen et al. 2021" and "Lee et al. 2016," which report more substantial effects. Notably, "Fang et al. 2025" presents a confidence interval that spans both negative and positive values, indicating a less certain effect compared to other studies. The overall test for heterogeneity is highly significant (p = 0.00), further underscoring the variability across studies. The test for the overall effect is also significant (p = 0.01), indicating that the treatment’s effect is statistically meaningful, although the degree of variability suggests caution in interpretation. In comparison to other studies in the field, this analysis provides a nuanced understanding of the treatment’s efficacy, highlighting both significant treatment effects and notable heterogeneity across the different studies included. Figure 3.

**Figure 3.**
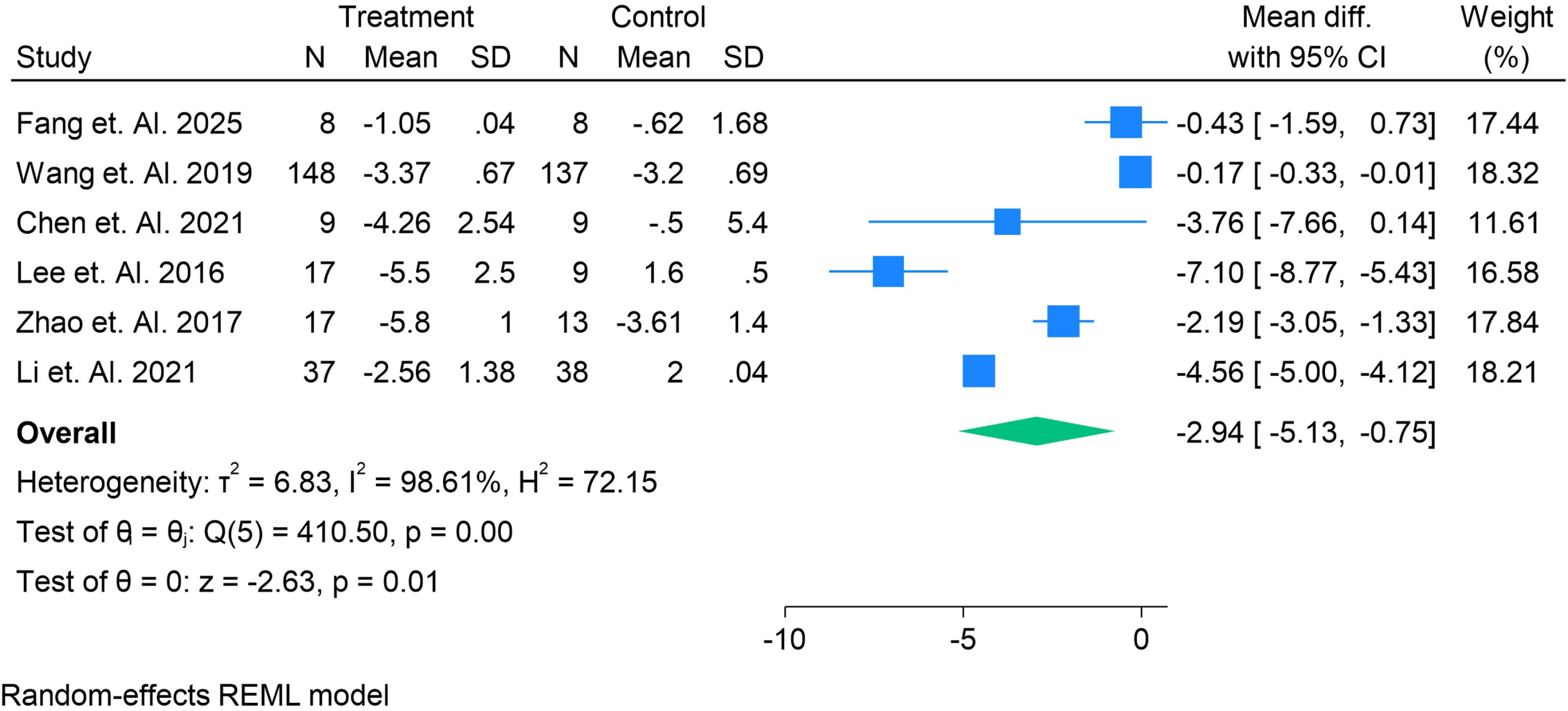
Meta-analysis of cognition after repetitive transcranial magnetic stimulation (rTMS) versus sham rTMS in ACOG

### Meta-Analysis of Mood, Global Impression, and Functional Performance After Repetitive Transcranial Magnetic Stimulation (rTMS) versus Sham rTMS

This meta-analysis examines the impact of repetitive transcranial magnetic stimulation (rTMS) on mood, global impression, and functional performance, measured by Instrumental Activities of Daily Living (IADL). The overall mean difference is -0.96 with a 95% confidence interval (CI) of [-6.21, 4.30], indicating no significant overall effect when comparing the treatment group with the sham rTMS control group. The heterogeneity (I² = 97.38%) is notably high, suggesting substantial variability across the studies included in the analysis, which could be attributed to differences in study populations, treatment regimens, or measurement tools.

In individual studies, "Ahmed et al. 2012" presents the largest mean difference of 6.80 (CI: [5.46, 8.14]), suggesting a significant improvement in functional performance following rTMS. Conversely, "Padala et al. 2020" reports a more substantial negative mean difference of -7.20 (CI: [-11.29, -3.11]), indicating a significant decline in functional performance with rTMS. Other studies, such as "Fang et al. 2025" and "Koch et al. 2022," show smaller but positive effects. The wide variation in results highlights the significant heterogeneity across the studies.

Despite these variations, the overall test for the effect (p = 0.72) shows no significant difference between the treatment and control groups, suggesting that rTMS may not produce a consistent impact on mood and functional performance across different study designs and populations. Further research with more standardized methods may help clarify the role of rTMS in improving functional performance and mood. Figure 4.

**Figure 4.**
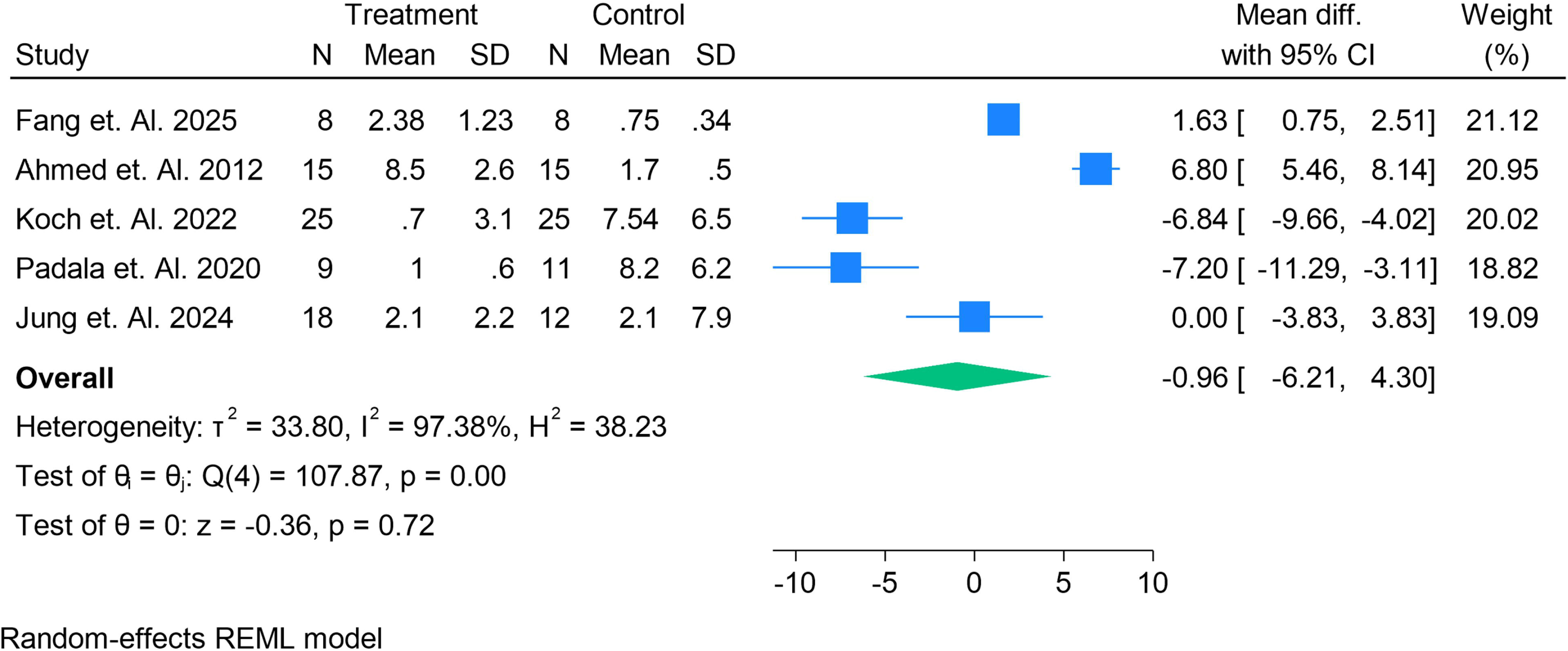
Meta-analysis of mood, global impression, function after repetitive transcranial magnetic stimulation (rTMS) versus sham rTMS. Change in functional performance as measured by IADL.

### Meta-Analysis of Adverse Events (Headache) after Repetitive Transcranial Magnetic Stimulation (rTMS) versus Sham rTMS

The forest plot presented in this meta-analysis investigates the incidence of adverse events like headache following repetitive transcranial magnetic stimulation (rTMS) compared to sham rTMS. The overall log risk ratio is -0.31 with a 95% confidence interval (CI) of [-0.84, 0.22], indicating no significant difference in the risk of headaches between the treatment and control groups. The heterogeneity is very low (I² = 3.20%), suggesting that the studies included are relatively consistent in their findings.

The individual studies show varying effects: "Koch et al. 2025" reports a significant positive log risk ratio of 0.85 (CI: [-1.34, 3.04]), suggesting a higher incidence of headaches in the treatment group compared to the control. In contrast, "Zhao et al. 2017" and "Padala et al. 2020" report negative log risk ratios (-2.43 and -0.29, respectively), indicating a reduced incidence of headaches in the treatment group compared to the sham. Other studies like "Sabagh et al. 2019" and "Lee et al. 2016" also show minimal differences between groups, but none of the confidence intervals exclude zero, supporting the overall finding of no significant difference.

The lack of statistical significance in the overall analysis (p = 0.25) and the narrow confidence interval suggest that rTMS does not consistently lead to increased adverse events like headaches when compared to sham treatment. This is further reinforced by the low heterogeneity, suggesting that the results across studies are largely in agreement despite some variation in individual findings. Figure 5.

**Figure 5.**
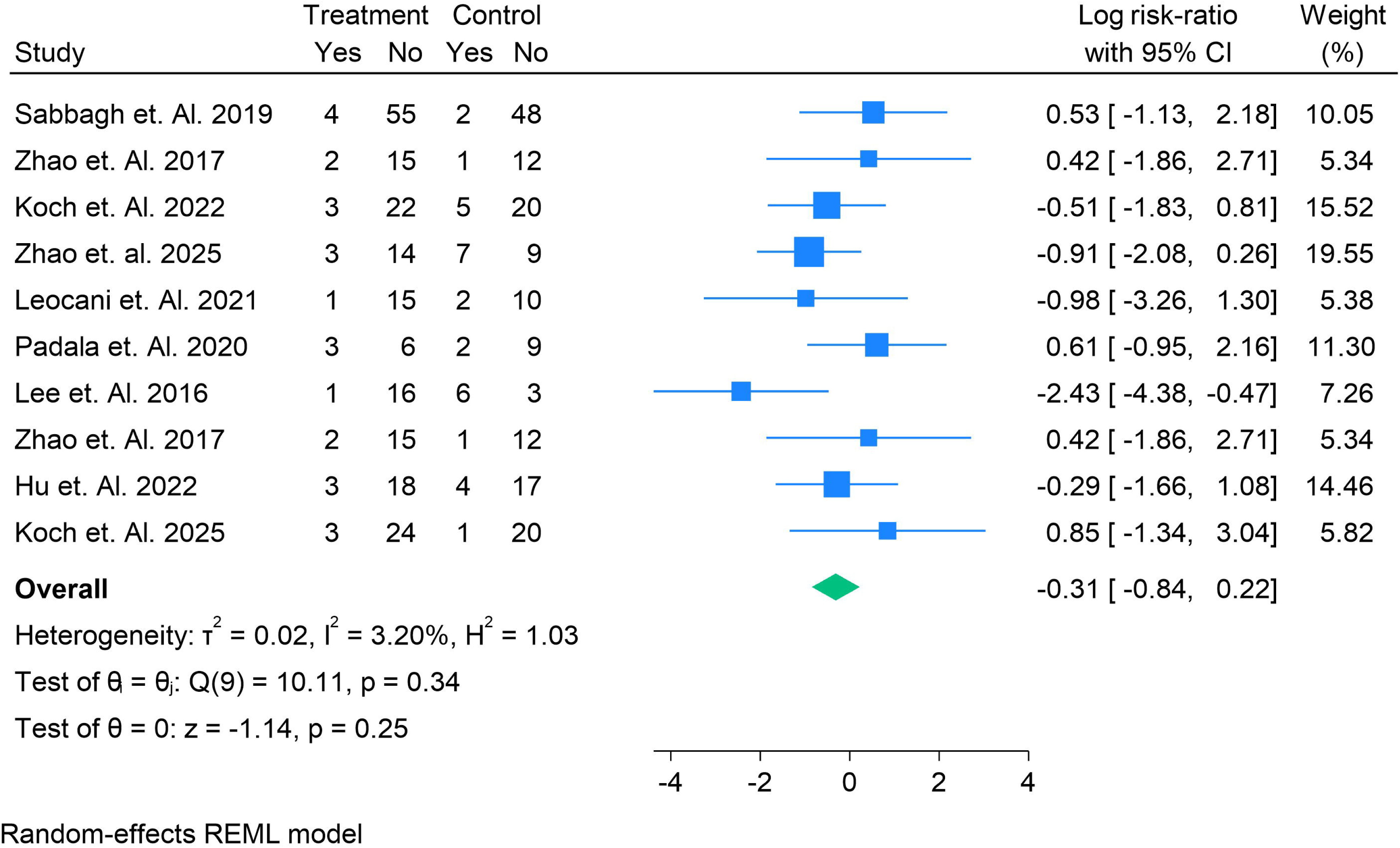
Meta-analysis of Adverse Events like Headache after repetitive transcranial magnetic stimulation (rTMS) versus sham rTMS.

### Meta-Analysis of Adverse Events (Scalp Pain) after Repetitive Transcranial Magnetic Stimulation (rTMS) versus Sham rTMS

This meta-analysis explores the incidence of scalp pain as an adverse event following repetitive transcranial magnetic stimulation (rTMS), comparing the treatment group to the sham rTMS control group. The overall log risk ratio is 0.40 with a 95% confidence interval (CI) of [-0.42, 1.21], indicating no significant difference in the risk of scalp pain between the two groups. The heterogeneity (I² = 21.32%) is relatively low, suggesting that the studies are more consistent in their findings.

The individual studies show mixed results: "Sabbagh et al. 2019" reports a log risk ratio of -0.17 (CI: [-2.09, 1.76]), suggesting no meaningful difference in scalp pain incidence between the treatment and control groups. However, "Koch et al. 2022" shows a positive log risk ratio of 1.39 (CI: [-0.73, 3.51]), indicating an increased risk of scalp pain in the treatment group. In contrast, "Saito et al. 2022" and "Padala et al. 2020" report negative log risk ratios (-0.29 and 1.81, respectively), suggesting a lower incidence of scalp pain with rTMS compared to the sham group.

Despite the variation across studies, the overall test for effect (p = 0.34) reveals no significant difference between the treatment and control groups. The narrow confidence interval further supports the lack of a substantial difference in scalp pain incidence following rTMS. This analysis suggests that rTMS does not significantly increase the risk of scalp pain, although individual study results may vary depending on factors such as treatment parameters and patient populations. Figure 6.

**Figure 6.**
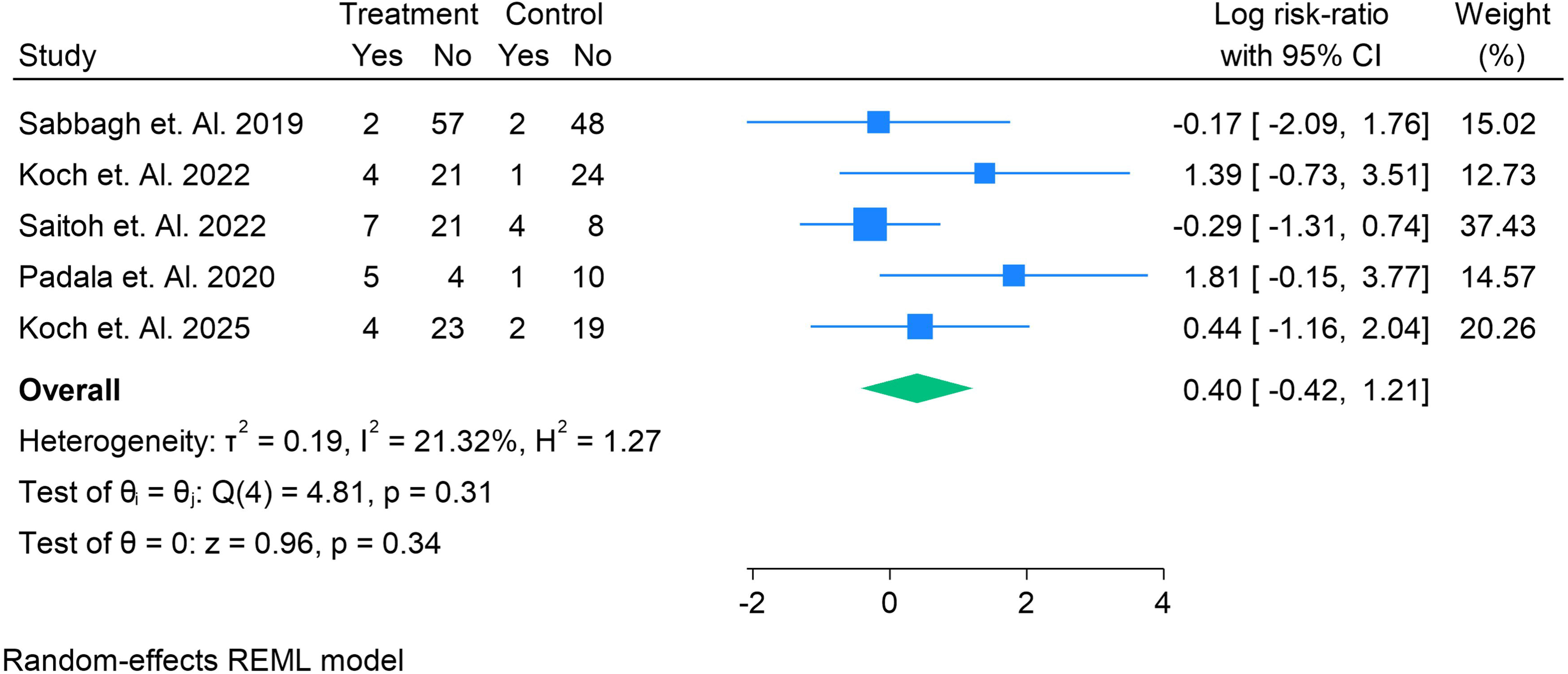
Meta-analysis of Adverse Events like Scalp Pain after repetitive transcranial magnetic stimulation (rTMS) versus sham rTMS.

## Discussion

Alzheimer’s disease (AD) is a progressive neurodegenerative disorder characterized by a decline in cognitive functions, including memory, executive function, and language, alongside significant changes in behavior and mood. Given the lack of disease-modifying pharmacological treatments, non-invasive therapies such as repetitive transcranial magnetic stimulation (rTMS) have garnered considerable interest in recent years for their potential to alleviate cognitive decline and improve other aspects of the disease. This systematic review and meta-analysis aimed to evaluate the efficacy of rTMS on cognitive function, mood, global impression, and functional abilities in individuals with Alzheimer’s disease, addressing an important gap in the current literature.

The results from this meta-analysis provide insights into the varying effects of rTMS across different outcome measures. While the overall effect of rTMS on cognition, assessed through the Mini-Mental State Examination (MMSE) and the Alzheimer’s Cognitive Scale (ADAS-cog), showed mixed results, with some studies indicating modest benefits and others revealing negligible or negative effects, it is clear that the heterogeneity across studies plays a major role in the observed variability. For example, the overall mean difference in cognition on the MMSE was -0.48 (95% CI [-1.41, 0.46]), which suggests a lack of significant difference between rTMS and sham rTMS. Similarly, the ADAS-cog scale also revealed a significant treatment effect in favor of rTMS, though the high heterogeneity (I² = 98.61%) in the analysis indicates substantial variability in effect sizes across studies. This suggests that the true effect of rTMS might vary across studies due to differences in patient characteristics, disease severity, and rTMS protocols.

One notable aspect of this meta-analysis is the significant variability between the studies, which is consistent with findings from previous systematic reviews and meta-analyses on rTMS in Alzheimer’s disease. A recent meta-analysis by Loo et al. (2018) examined the efficacy of rTMS in Alzheimer’s disease and found that while rTMS showed small to moderate effects on cognitive function, the effects were inconsistent across trials. Loo et al. also highlighted the importance of rTMS parameters (such as frequency and target brain region) and participant characteristics, including the severity of Alzheimer’s disease, in influencing the treatment outcomes [49]. Our study also found variability in the treatment effects across studies, reinforcing the notion that rTMS may be more effective in certain subgroups of patients, particularly those in the early stages of the disease.

In terms of mood and functional outcomes, this meta-analysis did not show a significant overall effect of rTMS compared to sham treatment, as the mean difference in mood, global impression, and functional performance was -0.96 (95% CI [-6.21, 4.30]). These findings align with those of other studies, such as the network meta-analysis by Jiang et al. (2021), which reported no significant impact of rTMS on functional performance or global impression in AD patients. However, individual studies within our meta-analysis, demonstrated substantial variation, with some studies showing significant improvement in functional performance and others showing deterioration [50]. The wide range of results further emphasizes the challenge of interpreting the clinical effectiveness of rTMS in AD.

The meta-analysis on adverse events, specifically headaches and scalp pain, also suggests that rTMS is relatively well-tolerated compared to sham treatments. The incidence of adverse events such as headaches and scalp pain was low and did not differ significantly between the rTMS and sham groups, which is consistent with the findings from other meta-analyses in this area. For instance, a meta-analysis by Ridding and Ziemann (2018) concluded that rTMS is a safe intervention with a low risk of side effects. Similarly, a recent study by Rossi et al. (2020) found that while some mild adverse effects, such as headaches, were reported, these were generally transient and did not lead to treatment discontinuation. Our analysis also found low heterogeneity in adverse events, reinforcing the safety profile of rTMS in AD [51].

However, several limitations should be considered when interpreting the results of this meta-analysis. First, the significant heterogeneity observed in the cognitive outcomes (I² = 99.42%) suggests that the studies included in this analysis may not be directly comparable. Variations in rTMS protocols, including stimulation frequency, intensity, and number of sessions, as well as differences in patient populations, could contribute to this heterogeneity. For example, the study by Zhao et al. (2017) reported a smaller effect of rTMS on cognitive function, which could be attributed to the differences in the treatment regimen or the inclusion of a more heterogeneous sample of patients [51]. In contrast, studies like Ahmed et al. (2012) and Koch et al. (2025) reported more substantial effects, suggesting that certain protocols or patient characteristics may optimize the benefits of rTMS.

Furthermore, the inclusion of fewer than 10 studies in some of the meta-analyses, such as the analysis of adverse events, limited the ability to assess publication bias. Larger, multi-center trials with standardized methodologies are needed to better understand the role of rTMS in AD. Additionally, most studies in this review focused primarily on short-term outcomes, and the long-term effects of rTMS remain unclear. Future research should explore the durability of rTMS-induced benefits over extended periods and the optimal parameters for treatment.

## Conclusion

In conclusion, while rTMS holds promise as a potential therapeutic intervention for Alzheimer’s disease, the findings from this meta-analysis indicate that its effects on cognition, mood, global impression, and functional performance remain inconsistent. The significant heterogeneity across studies highlights the need for more robust, standardized clinical trials to determine the most effective rTMS protocols and patient populations. Future studies should also explore the long-term benefits of rTMS and its potential role in combination with other therapeutic modalities.

## Data Availability

supplementary file

## Authors’ contributions

*All authors contributed equally to the manuscript and read and approved the final version of the manuscript*.

