## supplementary file for "Efficacy of Repetitive Transcranial Magnetic Stimulation in Alzheimer’s Disease: A Systematic Review and Meta-Analysis of Cognitive, Mood, and Functional Outcomes"

Search String

Table S1. Demographics and Summary table.

| **Ref No.** | **Author and Year** | **Country** | **Total Sample** | **Male** | **Female** | **Mean Age** | **Follow up time** | **Treatment Name** | **Treatment Sample** | **Control Name** | **Control Sample** | **Stimulation target Place** | **Stimulation Protocol** | **GRADE** | **Main finding** |
| --- | --- | --- | --- | --- | --- | --- | --- | --- | --- | --- | --- | --- | --- | --- | --- |
| 14 | Fang et. Al. 2025 | China | 16 | 5 | 11 | 66.38 | 1 | rtMS | 8 | Sham | 8 | Left dorsolateral prefrontal cortex (DLPFC) and left lateral temporal lobe (LTL) | 10 Hz stimulation, 100% resting motor threshold, 20 trains (5 s per train, 25-second intervals), 100 pulses per target area (10 min per target). | High | Rtms is better effiecient |
| 15 | Wang et. Al. 2019 | China | 240 | 120 | 120 | 71.2 | 12 | rtMS | 148 | Sham | 137 | Dorsolateral prefrontal cortex (DLPFC), Broca and Wernicke areas, superior temporal gyrus (STG), parietal somatosensory association cortex (pSAC). | Frequencies ranged from 1 Hz to 20 Hz, with session numbers varying from 2 to 30. | High | Rtms is better effiecient |
| 16 | Zhang et. Al. 2019 | China | 30 | 24 | 6 | 68.4 | 1 | rtMS | 15 | Sham | 15 | Left dorsolateral prefrontal cortex (DLPFC) and left lateral temporal lobe (LTL) | 10 Hz rTMS, 20 trains/day (1000 pulses/day), 5 days/week for 4 weeks. | High | Rtms is better effiecient |
| 17 | Sabbagh et. Al. 2019 | USA | 131 | 70 | 59 | 76.9 | 12 | rtMS | 59 | Sham | 50 | Broca’s area, Wernicke’s area, left/right dorsolateral prefrontal cortex, left/right inferior parietal lobule. | 10 Hz rTMS, 1300 pulses per session (20-pulse bursts), 30 sessions over 6 weeks. | High | Rtms is better effiecient |
| 18 | Vechhio et. Al. 2022 | Italy | 72 | 39 | 33 | 74.43 | 9.2 | rtMS | 30 | Sham | 16 | Broca’s area, R-DLPFC, L-DLPFC, Wernicke’s area, R-pSAC, L-pSAC. | 10 Hz rTMS (20–30 trains/day, 2 s/train, 20 pulses/train) for 6 weeks (30 sessions). | High | Rtms is better effiecient |
| 19 | Budak et. Al. 2022 | Turkey | 27 | 10 | 17 | 72 | 2 | rtMS | 19 | Sham | 8 | Bilateral dorsolateral 20prefrontal cortex(dlPFC). | 20 Hz rTMS, 3000 pulses per session (1500 per hemisphere), 10 sessions over 2 weeks. | High | Rtms is better effiecient |
| 20 | Zhao et. Al. 2017 | China | 30 | 10 | 20 | 70.8 | 1.5 | rtMS | 17 | Sham | 13 | Parietal (P3/P4) and posterior temporal (T5/T6) according to EEG 10-20 system. | Daily sessions for 6 weeks (1 session/day, 5 days/week, total 30 sessions). Each session included 10 min of rTMS (10 s of 20 Hz/train, 20 s intermediate/train). | High | Rtms is better effiecient |
| 21 | Leuchter et. Al. 2025 | USA | 54 | 27 | 27 | 63.5 | 1.25 | rtMS | 27 | Sham | 27 | Precuneus (a key hub in the default mode network). | 20 Hz rTMS, 1600 pulses per session, 100% of depth-corrected motor threshold, 16 sessions over 5 weeks. | High | Rtms is better effiecient |
| 22 | Ahmed et. Al. 2012 | Egypt | 45 | 16 | 29 | 68.4 | 3 | rtMS | 15 | Sham | 15 | Bilateral dorsolateral prefrontal cortex (DLPFC) | 20 Hz rTMS: 5 s, 20 trains, 25 s interstimulus interval, 2,000 pulses at 90% RMT | High | High-frequency (20 Hz) rTMS significantly improved cognitive function, daily living activities, and depression scores in mild-to-moderate Alzheimer's patients, with effects lasting up to 3 months. |
| 23 | Koch et. Al. 2022 | Italy | 50 | 24 | 26 | 73.7 | 24 | rtMS | 25 | Sham | 25 | Precuneus | 20 Hz, 40 trains of 2 seconds each, spaced by 28 seconds (total 1600 stimuli per session). Intensive phase: 5 sessions/week for 2 weeks. Maintenance phase: 1 session/week for 22 weeks. | High | Precuneus rTMS slowed cognitive and functional decline in Alzheimer’s disease patients over 24 weeks compared to sham treatment. |
| 24 | Chen et. Al. 2023 | China | 24 | 18 | 6 | 66.7 | 4 | rtMS | 18 | Sham | 18 | Left angular gyrus (MNI: -45, -67, 38) | 20 Hz frequency, 2 s stimulation, 28 s inter-interval, 100% resting motor threshold, 20 min duration, 1600 pulses per session, 5 sessions/week for 4 weeks. | High | Neuro-navigated rTMS targeting the left angular gyrus improved cognitive function in AD patients, with baseline DMN connectivity predicting treatment response. |
| 25 | Cotelli et. Al. 2008 | Italy | 47 | 24 | 24 | 75 | 12 | rtMS | 24 | Sham | 24 | Left and right dorsolateral prefrontal cortex (dlPFC, Brodmann area 8) | Online high-frequency rTMS (20 Hz for 500 ms at 90% resting motor threshold) | High | High-frequency rTMS to dlPFC improved naming performance in AD patients, with action naming improved in mild AD and both action/object naming improved in moderate-severe AD. |
| 26 | Tao et. Al. 2022 | China | 46 | 21 | 25 | 67 | 1.5 | rtMS | 23 | Sham | 23 | Left dorsolateral prefrontal cortex (DLPFC) | 20 Hz, 100% resting motor threshold intensity, 2 s stimulation followed by 25 s gap, 1760 pulses per session, 5 sessions/week for 6 weeks | Moderate | rTMS decreased serum Aβ levels and increased p75ECD levels in Alzheimer’s patients, correlating with improved cognitive function. |
| 27 | Jia et. Al. 2021 | China | 69 | 21 | 48 | 71.41 | 2 | rtMS | 35 | Sham | 34 | Left parietal cortex (highest functional connectivity to hippocampus). | 10 Hz rTMS, 100–110% motor threshold, 800 pulses per session, 10 sessions over 2 weeks. | Moderate | Precision rTMS over the left parietal cortex significantly improved memory and cognitive function in Alzheimer’s disease patients compared to sham treatment. |
| 28 | Chen et. Al. 2021 | China | 18 | 7 | 11 | 70.3 | 6 | rtMS | 9 | Sham | 9 | Left dorsolateral prefrontal cortex (L-DLPFC) | 20 Hz, 110% motor threshold, 1 s stimulation, 29 s interval, 1200 pulses per session, twice daily for 14 days. | High | Preliminary data suggest non-APOE4 carriers show better improvement in MoCA scores after rTMS compared to APOE4 carriers, indicating genotype-specific treatment responses. |
| 29 | Saitoh et. Al. 2022 | Japan | 42 | 15 | 25 | 76.4 | 6.5 | rtMS | 28 | Sham | 12 | Bilateral dorsolateral prefrontal cortex (DLPFC). | 15 trains bilaterally (40 pulses/train at 10 Hz; intertrain interval, 26 s). | High | rTMS at 120% RMT showed significant improvement in ADAS-Cog for patients with MMSE ≥ 15 compared to sham, with no serious adverse events. |
| 30 | Zhang et. Al. 2023 | China | 35 | 21 | 14 | 84.8 | 3 | rtMS | 18 | Sham | 17 | Left dorsolateral prefrontal cortex (DLPFC) | 10 Hz, 60 sessions over 3 months, 100% motor threshold (MT) | High | rTMS significantly improved cognitive performance (SIB), reduced psychiatric symptoms (NPI), and improved clinician’s global impression (CIBIC-Plus) in moderate-to-severe AD patients. |
| 31 | Zhao et. al. 2025 | China | 33 | 19 | 14 | 73.6 | 1 | rtMS | 17 | Sham | 16 | Bilateral dorsolateral prefrontal cortex (DLPFC). | 1-Hz stimulation to the right DLPFC and 10-Hz stimulation to the left DLPFC, 20 sessions over 4 weeks. | High | rTMS significantly improved depressive symptoms in Alzheimer's disease patients, with reduced prefrontal activation observed via fNIRS. |
| 32 | Leocani et. Al. 2021 | Italy | 28 | 14 | 14 | 70.9 | 4 | rtMS | 16 | Sham | 12 | Bilateral frontal-parietal-temporal regions; 10 Hz, 840 stimuli/session (42 trains of 20 stimuli, 22s intervals), 120% RMT, 3 sessions/week for 4 weeks, then 1 session/week for 4 weeks. | Not explicitly stated; overall baseline MMSE was 16.9 (SD 5.5). | High | rTMS with H-coil is feasible and safe, with transient cognitive improvement in AD patients. |
| 33 | Qin et. Al. 2023 | China | 21 | 5 | 16 | 65.6 | 1 | rtMS | 11 | Sham | 10 | Left dorsolateral prefrontal cortex (DLPFC) and left lateral temporal lobe (LTL). | 10 Hz rTMS, 20 min per day for 4 weeks, 1,000 pulses per session. | High | High-frequency rTMS combined with cognitive training improved cognitive function and activities of daily living in mild-to-moderate AD patients, with immediate CBF reduction in the precuneus and long-term CBF increase in the left parahippocampus. |
| 34 | Padala et. Al. 2020 | USA | 20 | 18 | 5 | 77.3 | 3 | rtMS | 9 | Sham | 11 | Left dorsolateral prefrontal cortex (DLPFC). | 10 Hz, 3000 pulses per session, 120% motor threshold, 4-second train duration, 26-second inter-train interval, for 20 consecutive weekdays. | High | rTMS significantly improved apathy, cognition (3MS), and daily function (IADL) in Alzheimer’s patients compared to sham treatment, with effects durable at 12 weeks. |
| 35 | Lee et. Al. 2016 | Korea | 26 | 11 | 15 | 71 | 3 | rtMS | 17 | Sham | 9 | Six cortical areas (bilateral dorsolateral prefrontal cortex, Broca’s area, Wernicke’s area, bilateral parietal somatosensory association cortex). | 10 Hz, 90–110% motor threshold intensity, 20 trains/day (2 s/train, 20 pulses/train), 5 days/week for 6 weeks (total 30 sessions). | High | rTMS-COG improved cognitive function (ADAS-cog, MMSE) in mild Alzheimer’s patients, particularly in memory and language domains, with sustained effects at 6 weeks post-treatment. |
| 36 | Jung et. Al. 2024 | South Korea | 30 | 18 | 12 | 69.8 | 2 | rtMS | 18 | Sham | 12 | Left parietal area functionally connected to the hippocampus (based on fMRI). | 20 sessions over 4 weeks (40 trains/day, 20 Hz for 2 seconds, 1600 pulses/day). | High | Personalized hippocampal network-targeted rTMS significantly improved cognition and functional performance in Alzheimer’s disease patients compared to sham stimulation. |
| 37 | Wei et. Al. 2024 | China | 120 | 60 | 60 | 65 | 12 | rtMS | 60 | Sham | 60 | Left parietal area functionally connected to the hippocampus (based on fMRI). | | High | rTMS improved cognitive function (memory, attention, executive function) in patients with Alzheimer’s disease (AD) and cerebral small vessel disease (CSVD), with AD patients showing greater memory/executive improvements and CSVD patients showing better attentional gains. |
| 38 | Moussavi et. Al. 2024 | Canada | 135 | 85 | 71 | 74 | 6 | rtMS | 105 | Sham | 51 | Bilateral dorsolateral prefrontal cortex (DLPFC) | 20 Hz, 30 pulses/train, 25 trains, 10-s intertrain interval | High | Active rTMS was not superior to sham rTMS for improving cognition in mild-to-moderate Alzheimer’s disease. |
| 39 | Yuan et. Al. 2021 | China | 24 | 11 | 13 | 65.08 | 1 | rtMS | 12 | Sham | 12 | Left dorsolateral prefrontal cortex (DLPFC). | 10 Hz rTMS (2s on, 8s off, 20 reps at 80% motor threshold, 400 pulses/session), delivered 5x/week for 4 weeks to the left DLPFC. | High | High-frequency rTMS over the left DLPFC improved cognitive function and altered spontaneous brain activity in cognitive-related areas in patients with amnestic mild cognitive impairment (aMCI). |
| 40 | Padala et. Al. 2018 | USA | 9 | 8 | 1 | 65.5 | 2 | rtMS | 4 | Sham | 5 | Left dorsolateral prefrontal cortex (DLPFC); | 10Hz, 3000 pulses/session, 120% motor threshold, 5 days/week for 2 weeks. | High | rTMS significantly improved apathy and cognition in MCI patients compared to sham treatment. |
| 41 | Zhao et. Al. 2017 | China | 30 | 15 | 15 | 70.8 | 1.5 | rtMS | 17 | Sham | 13 | Parietal P3/P4 and posterior temporal T5/T6 (according to EEG 10-20 system) | Daily sessions for 6 weeks (1 session/day, 5 days/week, total 30 sessions). Each session included 10 min of rTMS (10 s of 20 Hz/train, 20s intermediate/train). | Moderate | Repetitive transcranial magnetic stimulation (rTMS) improves cognitive function, memory, and language in Alzheimer’s disease patients, especially in the mild stage. |
| 42 | Li et. Al. 2021 | China | 75 | 44 | 17 | 31 | 3 | rtMS | 37 | Sham | 38 | Left DLPFC (MNI coordinates: -44, 40, 29). | 20 Hz stimulation, 1-s pulse train duration, 10-s intertrain intervals, 100 trains/day (2000 pulses/day), 30 sessions over 6 weeks. | Moderate | Cortical LTP-like plasticity correlated with cognitive improvement in AD patients after rTMS treatment. |
| 43 | Yao et. Al. 2022 | China | 27 | 14 | 13 | 65.4 | 12 | rtMS | 15 | Sham | 15 | Bilateral cerebellum Crus II. | 5 Hz rTMS, 2000 pulses per session, 20 sessions over 4 weeks (5 days/week). | High | 5 Hz rTMS over the bilateral cerebellum significantly improved multi-domain cognitive functions in AD patients by modulating cerebello-cerebral connectivity. |
| 44 | Hu et. Al. 2022 | China | 84 | 38 | 46 | 77.16 | 2 | rtMS | 21 | Sham | 21 | Bilateral angular gyrus (AG, P5/P6 electrode site) | Simultaneous 40-Hz rTMS (90% RMT) and 2-mA tDCS (anode over AG, cathode over contralateral prefrontal area), 3 times/week for 4 weeks. | High | Simultaneous rTMS–tDCS over bilateral AG significantly improved neuropsychiatric symptoms, cognition, and sleep quality in moderate AD patients compared to sham or unimodal stimulation. |
| 45 | Kumar et. Al. 2023 | Canada | 32 | 16 | 16 | 76.4 | 14 | rtMS | 16 | Sham | 16 | Left dorsolateral prefrontal cortex (DLPFC). | 180 pulses at 0.1 Hz, interstimulus interval = 25 ms. | High | The study did not show significant differences between active and control rPAS on DLPFC plasticity or working memory, but post hoc analyses suggested acute improvements in the active rPAS group. |
| 46 | Rabey et. Al. 2013 | Israel | 15 | 10 | 5 | 72.6 | 4.5 | rtMS | 7 | Sham | 8 | Six cortical brain regions (R-dIPFC, L-dIPFC, Broca, Wernicke, R-pSAC, L-pSAC). | 10 Hz rTMS, 20 trains of 2 s each per brain region, totaling 1,300 pulses/day. | High | rTMS-COG is a safe and effective therapy for improving cognitive function in Alzheimer’s disease patients. |
| 47 | Mencarelli et. Al. 2024 | Italy | 16 | 8 | 8 | 62 | 6 | rtMS | 8 | Sham | 8 | Precuneus (PC) | 10 daily sessions (20 Hz, 40 trains of 2 s each, spaced by 28 s, total 1600 stimuli per session). | High | PC-rTMS preserved grey matter integrity in the precuneus and increased functional connectivity within the DMN in Alzheimer’s disease patients after 24 weeks of treatment. |
| 48 | Koch et. Al. 2025 | Italy | 48 | 21 | 27 | 72.8 | 12 | rtMS | 27 | Sham | 21 | Precuneus (PC) | 10 daily sessions (20 Hz, 40 trains of 2 s each, spaced by 28 s, total 1600 stimuli per session). | High | PC-rTMS over 52 weeks slowed cognitive decline, preserved daily living activities, and reduced behavioral disturbances in mild-to-moderate AD patients compared to sham. |

Figure S1. Risk of Bias

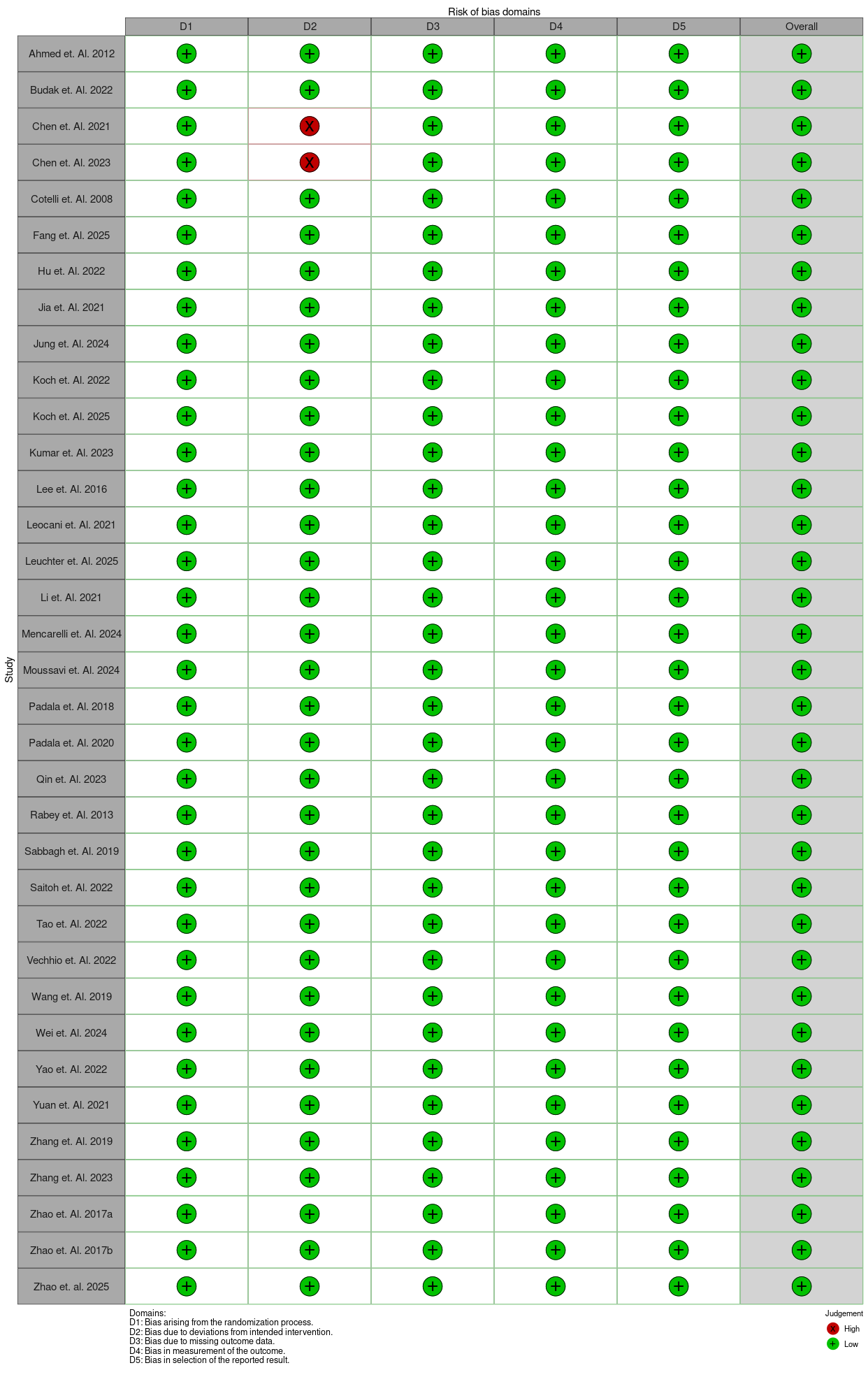
